# A Transdiagnostic fMRI Dataset With 300+ Deeply Phenotyped Subjects Across Resting and Task States

**DOI:** 10.1101/2025.07.15.25331595

**Authors:** Anja Samardzija, Xilin Shen, Abigail S. Greene, Saloni Mehta, Santino Iannone, Alexander J Simon, Flor Parra, Wenjing Luo, Jagriti Arora, Fuyuze Tokoglu, C. Alice Hahn, Scott W. Woods, Gerard Sanacora, Rachel Katz, Vinod Srihari, Marisa N. Spann, Dustin Scheinost, R. Todd Constable

## Abstract

YaleNeuroConnect is a human functional MRI (fMRI) dataset collected at Yale University that includes functional MRI data (and the respective functional connectomes) obtained under resting-state and six task conditions. There are 302 diagnostically and demographically diverse subjects, each with extensive neuropsychological testing and symptom inventories obtained outside of the MRI. Prior studies have shown that stronger predictive models relating the brain to external measures can be built with connectivity data obtained during continuous performance tasks instead of the more common resting-state. The tasks here were selected to exercise the brain across various cognitive domains. For each subject, 48 minutes of fMRI data and high-resolution 3D brain volumes were obtained. The fMRI data, along with the deep phenotyping data in a diverse subject pool, allow studies of brain parcellation under different conditions, the relationship between cognitive and clinical measures, identification of circuits supporting external measures, and data for the development of brain-based tests. The transdiagnostic nature of the sample allows a sufficient range of symptom scores to test the principles of the Research Domain Criteria framework.

## Background & Summary

This functional magnetic resonance imaging (fMRI) dataset is rich along dimensions not commonly found in current fMRI datasets. First, this is a truly trans-diagnostic sample of individuals. The subjects span a wide range of mental health profiles, including subjects that endorse no symptoms on symptom inventories to subjects that check multiple symptoms. While studies often require diagnostic specificity (e.g., major depressive disorder without comorbid conditions), this dataset reflects real-world comorbidities—a significant strength. Second, the subjects are deeply phenotyped. In addition to the imaging protocol, subjects underwent a two-hour battery of standardized cognitive testing and clinical symptom score checklists, revealing a comprehensive psychological profile of each individual. Third, the focus is on functional connectivity data, and as such, this dataset contains not only 16 minutes of resting-state data per subject but also functional MRI data collected across six different continuous performance tasks aimed at altering brain state, with study design optimized for the calculation of functional connectivity. These tasks include a Card Guessing^1-3^ task, a Reading the Mind in the Eyes^4^ task, a Gradual-Onset Continuous Performance (gradCPT)^5^ task, a Movie Watching task, a n-back^6-8^ task, and a Stop-Signal^9,10^ task (SST). These tasks are designed to reconfigure the brain along different cognitive axes. The fMRI data was used to generate the functional connectomes by segmenting the brain into nodes (brain regions defined by an atlas) using the predefined function-based Shen268^11^ brain atlas and Shen368^12^ brain atlas, and quantifying the functional connectivity as the pairwise correlations between the mean time courses of voxels in each node.

This dataset was collected with the intention of its use in brain-behavior modeling to reveal the networks associated with cognitive and clinical systems, to understand node reconfiguration as a function of brain state^13-15^, to observe how predictive modeling is influenced by task^16-19^, as well as to develop a brain-based approach to designing new test instruments. By understanding the link between behavior/clinical symptoms and the functional organization of the individual’s brain, we may be able to extract functional phenotypes that aid in the clinical characterization of individual patients^20^.

Models that only work on specific patient groups or control subjects offer limited clinical utility. Instead, modeling methods that are designed to generalize across multiple behaviors and diagnostic groups are needed^7^. There is considerable work demonstrating links between cognitive alterations and clinical symptoms^21-24^. Cognitive deficits may appear before the first psychotic episode in schizophrenia^25^ and have been linked to a range of mental disorders^26^. This dataset consists of healthy controls and patients with major depressive disorder (MDD), prodromal psychosis, first episode psychosis, anxiety disorders, obsessive-compulsive disorder (OCD), post-traumatic stress disorder (PTSD), bipolar disorder, etc. (Figure 1 and Table 1). The clinical and demographic details of the dataset are listed in Tables 1-2 and Figures 1-2. The collection of this transdiagnostic dataset was motivated by the need to develop models directly related to behavioral variables, as well as to latent factors of clinical scores derived from brain-behavior models, to develop models related to transdiagnostic factors, such as internalizing and externalizing^8-10,27-34^. Such models then allow us to identify the relevant functional phenotypes that vary with these scores, placing patients on a continuum, and/or allowing clustering of patients into subgroups according to their functional phenotype as characterized by the brain-behavior models.

**Table 1.** Mental health diagnosis by sex.

| Diagnosis | Female | Male | Total |
| --- | --- | --- | --- |
| All | 170 | 132 | 302 |
| No-mental-health diagnosis | 80 | 60 | 140 |
| Major depressive episode (MDE) | 55 | 31 | 86 |
| Generalized anxiety disorder (GAD) | 48 | 21 | 69 |
| Manic episode | 0 | 3 | 3 |
| Bipolar disorder | 17 | 15 | 32 |
| Alcohol use disorder (AUD) | 12 | 15 | 27 |
| Substance use disorder (SUD) | 14 | 17 | 31 |
| Panic episode | 25 | 9 | 34 |
| Post-traumatic stress disorder (PTSD) | 18 | 8 | 26 |
| Social anxiety disorder (SAD) | 7 | 4 | 11 |
| Obsessive compulsive disorder (OCD) | 12 | 6 | 18 |
| Autism spectrum disorder (ASD) | 1 | 1 | 2 |
| Attention-deficit/ hyperactivity disorder (ADHD) | 17 | 16 | 33 |
| Agoraphobia | 9 | 4 | 13 |
| Psychotic episode/ schizophrenia | 9 | 18 | 27 |
| Borderline personality disorder (BPD) | 7 | 1 | 8 |

**Table 2.** Demographic and clinical information of participants. *μ*: mean, *σ*: standard deviation.

|  |  |
| --- | --- |
| **Age** | $\mu = 30.81$ $\sigma = 10.93$ |
| **Years of education** | $\mu = 15.85$, $\sigma = 3.13$ |
| **English as first language** | Yes = 259, No = 43 |
| **Stress** | $\mu = 22.78$, $\sigma = 9.62$ |
| **PSQI** | $\mu = 6.31$, $\sigma = 3.66$ |
| **Positive affect** | $\mu = 31.6$, $\sigma = 8.26$ |
| **Negative affect** | $\mu = 18.21$, $\sigma = 7.6$ |

**Figure 1.**
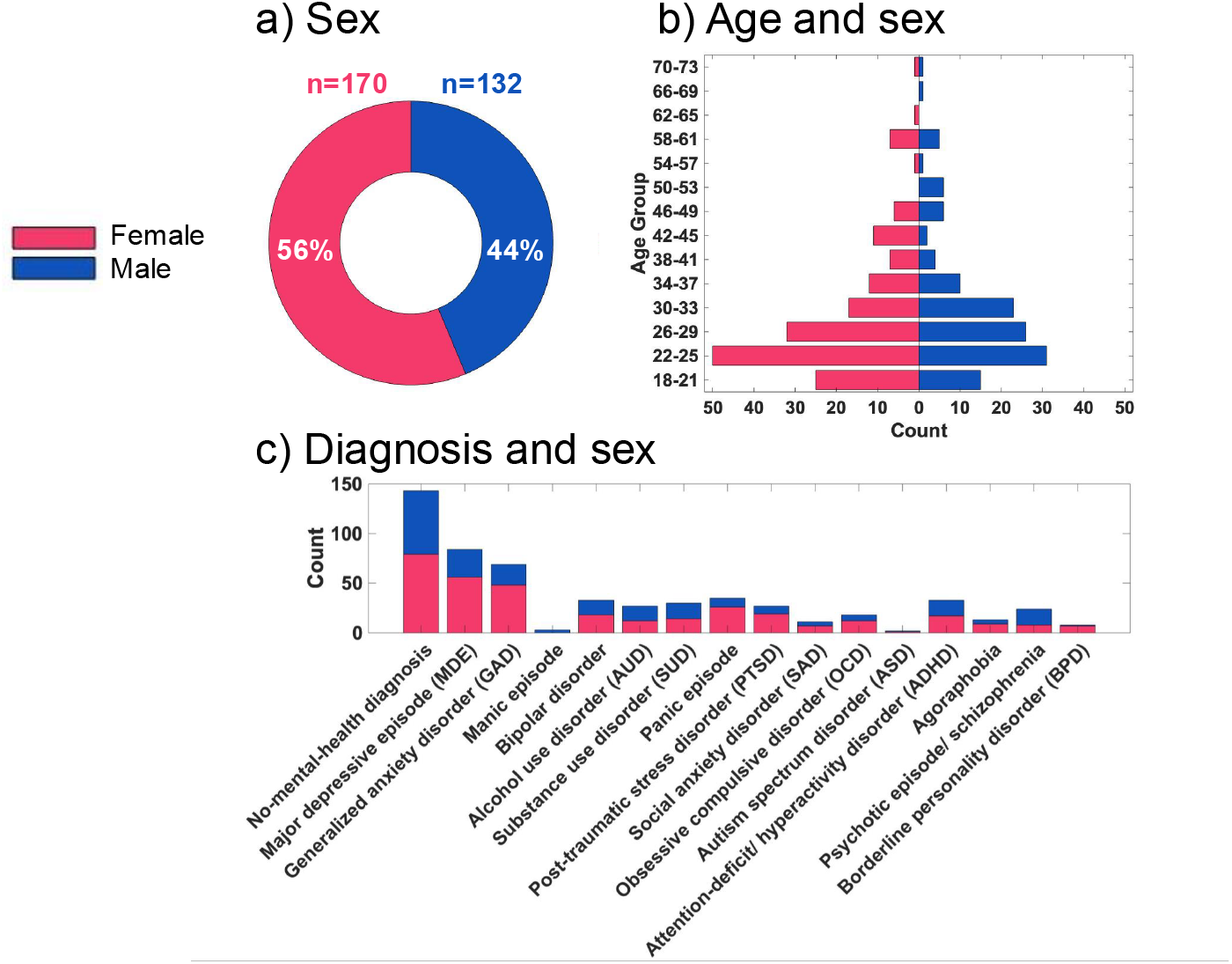
Dataset by sex. Pink depicts female participants; blue depicts male participants. a) Sex: n=170 of participants are Female (56%) and n=132 are Male (44%). b) Age and sex: count of participants by age group and sex. c) Diagnosis and sex: count of participants by mental health diagnosis and sex (non-exclusive comorbidities). Count of participants by mental health diagnosis and sex is listed in Table 1.

**Figure 2.**
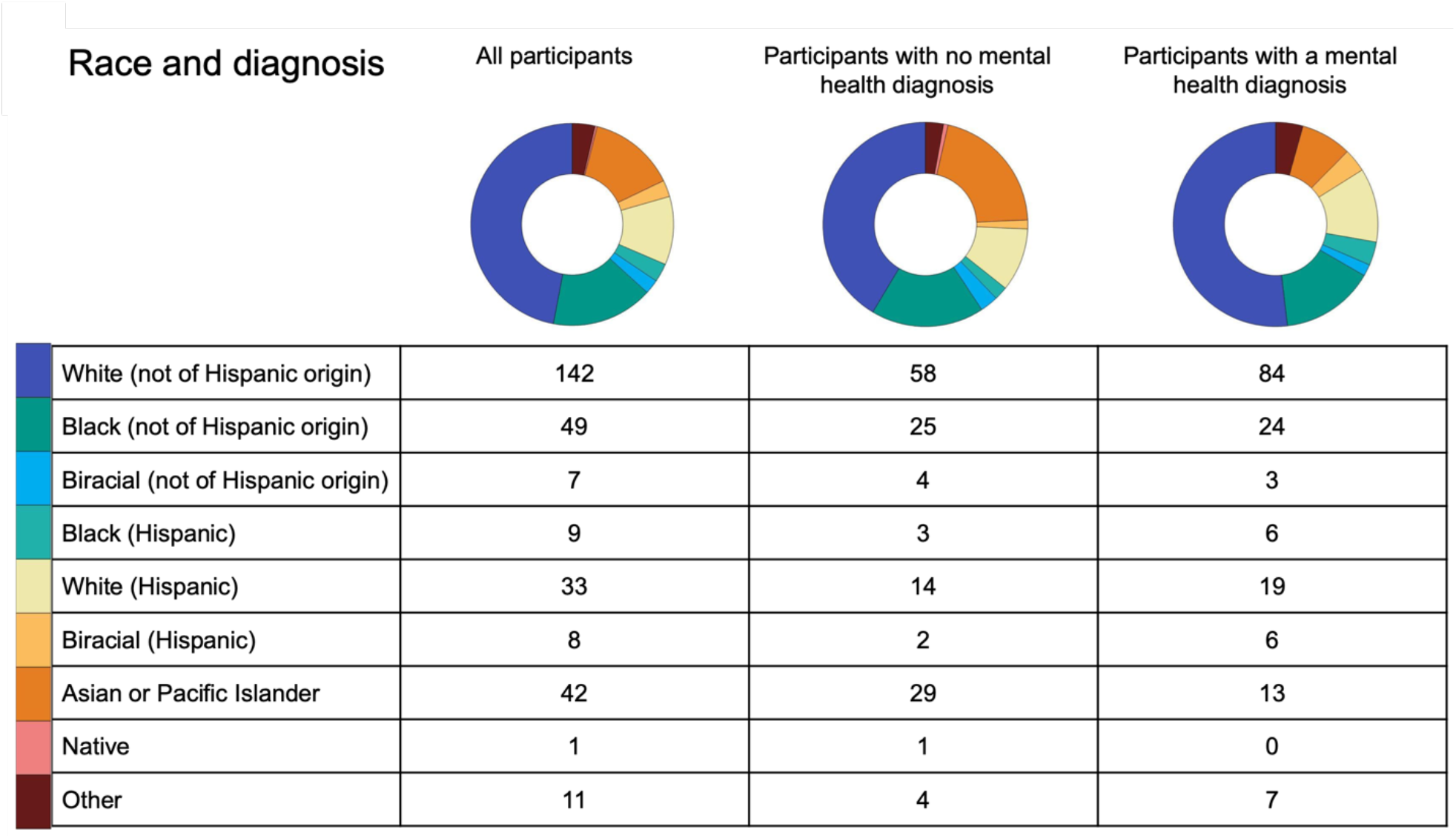
Race and diagnosis (n): all participants (second column), participants with no mental health diagnosis (third column), participants with a mental health diagnosis (fourth column). Participant race was self-reported. For rows 1–8, participants selected from predefined options; row 9 (“Other”) allowed for a written response. The responses to “Other” include: “Multiracial Hispanic”, “African”, “Half Asian-Indian”, “Brazilian”, “White (not of Hispanic origin) and Native”, “White (not of Hispanic origin) and Middle Eastern”, “White (not of Hispanic origin) and Asian or Pacific Islander”, “Hispanic”, “‘Blended heritage- Irish, Scottish, Malaysian, Indian, Khoisan”; the remaining “Other” responses were left blank.

To date, most functional brain-behavior modeling has been performed using resting state data (in which case the participant does not perform any task or activity while in the scanner). Individual functional connectomes look more similar to each other during tasks (where the participant performs a task while in the scanner) than during rest^35^, and the functional connectivity differences revealed by tasks lead to better predictive models that better identify the relevant neural circuits^36^. Thus, in conjunction with two resting-state runs, we collect six task runs. The series of six tasks, selected based on the *Behavioral assessment methods for RDoC constructs*^37^, was designed to enhance neurocognitive differences across individuals by directly tapping into specific fundamental cognitive processes. The six task scans were generated with the hypothesis that they would better reveal circuits associated with behavior and disease compared to resting-state data. Furthermore, through this multi-task approach, where functional connectivity matrices are formed from a series of cognitive tasks and resting state, the relevant connections that relate to latent factors of symptoms and behaviors will be better revealed compared to single-condition models^11^.

Part of the dataset has so far been used to show that brain-behavior models often reflect stereotypical profiles, and fail when applied to individuals who do not fit those profiles^38^. These results were replicated with two independent and publicly available datasets, the Human Connectome Project (HCP)^39^ and the UCLA Consortium for Neuropsychiatric Phenomics^40^. The results of this study demonstrated the limitations of applying a singular modeling approach to all subjects and highlighted that biased phenotypic measures limit the interpretability and utility of predictive models. A framework is presented to consider and address the impact of such bias so that predictive models reveal the brain circuits associated with phenotypes of interest, which will allow the exploration of individualized, cognitively specific neural targets for clinical intervention. The dataset has also been used in a study in which brain networks are predefined and the contribution of each brain network on widely used behavioral measures (and clinical scores) is evaluated^41^. This framework opens avenues of research for selecting the best outside-of-the-scanner test (based on brain data) for probing specific brain networks as well as guiding the development of new and better tests for probing brain networks of interest.

Similarly, future work using this dataset can explore the significance of different tasks in predicting behavioral and clinical measures. The broad diversity in diagnostic profiles among the participants along with the extensive range of behavioral and clinical scores collected alongside resting and task imaging data provides a solid foundation for conducting a wide variety of brain, behavior, and clinical studies using this dataset.

## Methods

### Participants

The dataset was collected at Yale School of Medicine, Magnetic Resonance Research Center, New Haven, Connecticut, USA between February 2018 and August 2024. A diverse participant sample was intentionally recruited, representing diagnostic variation, differing levels of symptom severity and mental health comorbidities, spanning a range of ages, and individuals from various racialized groups (Figures 1-2 and Tables 1-2).

Participants were recruited within the local community using widespread advertisement and through referrals from Yale clinics, including the Yale Depression Program, the PRIME research clinic, and the STEP clinic. We confirm that the participants are safe to undergo an MRI before scheduling them for the study. On the day of the study, participants fill out an MR safety form. Written informed consent for participation and data sharing in accordance with a protocol approved by the Yale Institutional Review Board (HIC #2000020891) is obtained from all participants. The study consists of five parts: 1) MRI scans, 2) self-report questionaries, 3) demographic and background information, 4) brief psychiatric interview, and 5) a battery of cognitive tests. The whole study takes approximately 2-3 hours. 1) The scanning takes approximately 90 minutes during which time anatomical scans and two resting fMRI runs and six task fMRI runs are run. 2) Following the scans, the participant is brought to the behavioral testing room where they complete a set of questionnaires regarding mood and sleep assessments (described in rows 2-8 of Table 3). 3) The participant fills out a diagnostic and background information form, which includes questions about mental health or neurological diagnoses, current medications, highest level of education, occupation, and annual household income. Study coordinators next conduct a 4) brief psychiatric interview (described in row 9 of Table 3) and 5) administer a battery of cognitive tests (described in Table 4).

**Table 3:** Outside-of-the-scanner self-report and interview-based psychological measures.

| **Measure** | **Sub-scales** | **Description** |
| --- | --- | --- |
| Behavior Rating Inventory of Executive Function – Adult (BRIEF-A)<sup>52</sup> | Inhibit, Self-monitor, Plan/organize, Shift, Initiate, Task Monitor, Emotional Control, Working Memory, Organization of Materials, Global Executive Composite, Behavioral Regulation Index, Metacognition Index, Negativity, Infrequency, Inconsistency | Self-report, 75 total items, raw and *T* score reported |
| Brief Symptom Inventory (BSI)<sup>53</sup> | Somatization, Obsession-Compulsion, Interpersonal Sensitivity, Depression, Anxiety, Hostility, Phobic Anxiety, Paranoid Ideation, Psychoticism, Additional Clinical Items | Self-report, 53 total items, raw and *T* score reported |
| Positive And Negative Affect Schedule – Expanded (PANAS-X)<sup>54</sup> | Negative Affect, Positive Affect, Fear, Hostility, Guilt, Sadness, Joviality, Self-assurance, Attentiveness, Shyness, Fatigue, Serenity, Surprise | Self-report, 60 total items, raw score reported |
| Adult Temperament Questionnaire (ATQ)<sup>55</sup> | Fear, Sadness, Discomfort, Frustration, Sociability, Positive Affect, High Intensity Pleasure, Attentional Control, Inhibitory Control, Activation Control, Neutral Perceptual Sensitivity, Affective Perceptual Sensitivity, Associative Sensitivity | Self-report, 77 total items, raw score reported |
| Interpersonal Reactivity Index (IRI)<sup>56</sup> | Perspective Taking, Empathetic Concern, Personal Distress, Fantasy | Self-report, 28 total items, raw score reported |
| Perceived Stress Scale (PSS)<sup>57</sup> | Stress Perception | Self-report, 10 total items, raw score reported |
| Pittsburgh Sleep Quality Index (PSQI)<sup>58</sup> | Quality, Latency, Duration, Habitual Efficiency, Disturbances, Medication, Daytime Dysfunction | Self-report, 19 total items, raw score reported |
| Mini International Neuropsychiatric Interview (MINI)<sup>59</sup> | Major Depressive Episode (MDE), Dysthymia, Suicidality, Hypomanic Episode, Panic Disorder, Agoraphobia, Social Phobia, Obsessive-Compulsive Disorder (OCD), Posttraumatic Stress Disorder (PTSD), Alcohol Use Disorder (AUD), Substance Use Disorder (SUD), Psychotic Disorders, Anorexia Nervosa, Bulimia Nervosa, Generalized Anxiety Disorder (GAD), Antisocial Personality Disorder | Interview, diagnoses reported with time frame (past, lifetime, recurrent, current) and severity/ context (if applicable) |

**Table 4:**
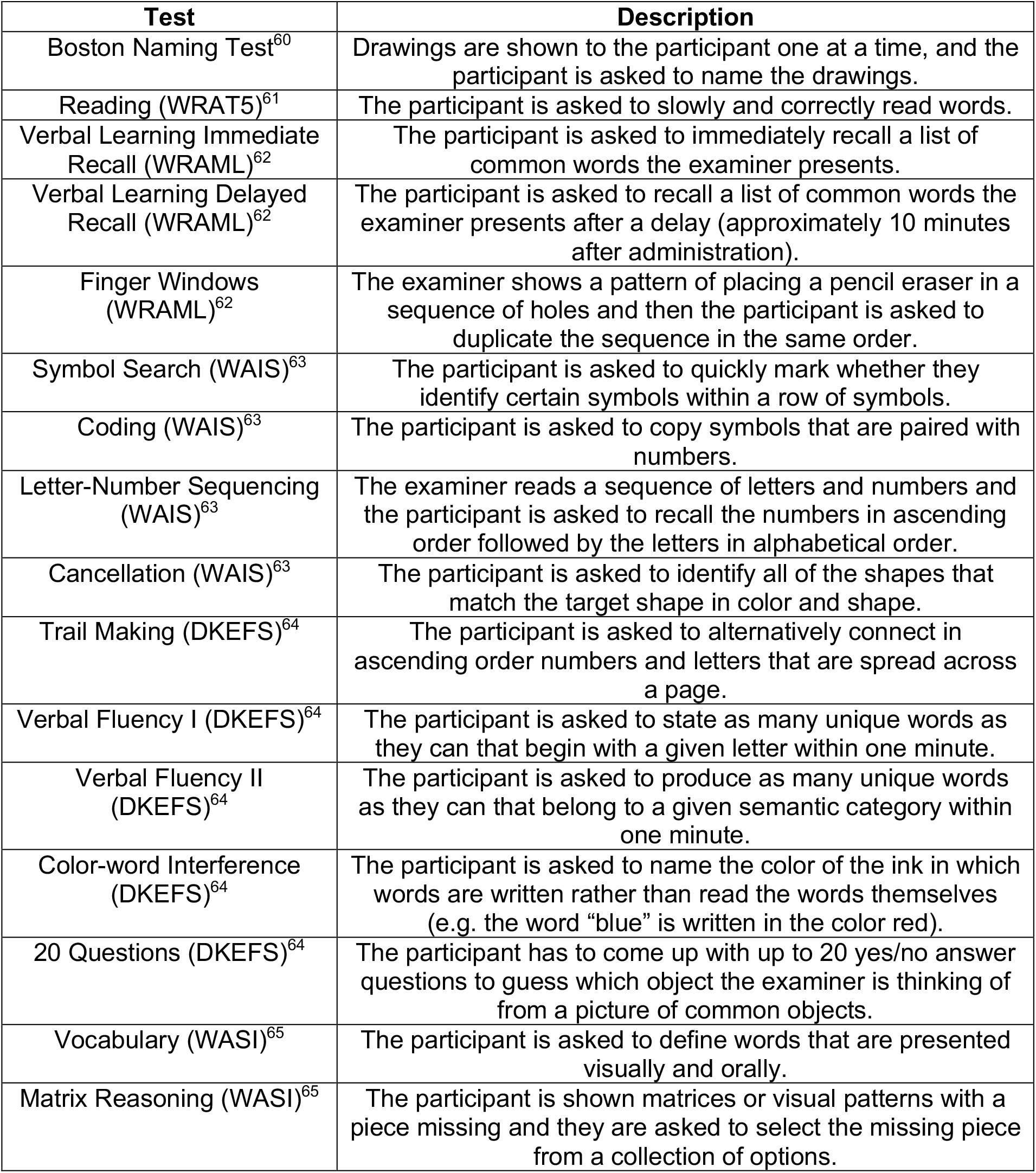
Outside-of-the-scanner cognitive measures.

| **Test** | **Description** |
| --- | --- |
| Boston Naming Test<sup>60</sup> | Drawings are shown to the participant one at a time, and the participant is asked to name the drawings. |
| Reading (WRAT5)<sup>61</sup> | The participant is asked to slowly and correctly read words. |
| Verbal Learning Immediate Recall (WRAML)<sup>62</sup> | The participant is asked to immediately recall a list of common words the examiner presents. |
| Verbal Learning Delayed Recall (WRAML)<sup>62</sup> | The participant is asked to recall a list of common words the examiner presents after a delay (approximately 10 minutes after administration). |
| Finger Windows (WRAML)<sup>62</sup> | The examiner shows a pattern of placing a pencil eraser in a sequence of holes and then the participant is asked to duplicate the sequence in the same order. |
| Symbol Search (WAIS)<sup>63</sup> | The participant is asked to quickly mark whether they identify certain symbols within a row of symbols. |
| Coding (WAIS)<sup>63</sup> | The participant is asked to copy symbols that are paired with numbers. |
| Letter-Number Sequencing (WAIS)<sup>63</sup> | The examiner reads a sequence of letters and numbers and the participant is asked to recall the numbers in ascending order followed by the letters in alphabetical order. |
| Cancellation (WAIS)<sup>63</sup> | The participant is asked to identify all of the shapes that match the target shape in color and shape. |
| Trail Making (DKEFS)<sup>64</sup> | The participant is asked to alternatively connect in ascending order numbers and letters that are spread across a page. |
| Verbal Fluency I (DKEFS)<sup>64</sup> | The participant is asked to state as many unique words as they can that begin with a given letter within one minute. |
| Verbal Fluency II (DKEFS)<sup>64</sup> | The participant is asked to produce as many unique words as they can that belong to a given semantic category within one minute. |
| Color-word Interference (DKEFS)<sup>64</sup> | The participant is asked to name the color of the ink in which words are written rather than read the words themselves (e.g. the word “blue” is written in the color red). |
| 20 Questions (DKEFS)<sup>64</sup> | The participant has to come up with up to 20 yes/no answer questions to guess which object the examiner is thinking of from a picture of common objects. |
| Vocabulary (WASI)<sup>65</sup> | The participant is asked to define words that are presented visually and orally. |
| Matrix Reasoning (WASI)<sup>65</sup> | The participant is shown matrices or visual patterns with a piece missing and they are asked to select the missing piece from a collection of options. |

### MRI Acquisition Protocol

The imaging data was collected at Yale on a 3T Siemens Prisma scanner with a 64-channel head coil. A high-resolution 3D anatomical scan (T1-weighted magnetization-prepared rapid acquisition with gradient-echo (MPRAGE) sequence [208 slices acquired in the sagittal plane, repetition time (TR)= 2,400 ms, echo time (TE) = 1.22 ms, flip angle = 8°, slice thickness = 1 mm, in-plane resolution = 1 mm × 1 mm]) was obtained for alignment to common space. The functional data were obtained using a multiband gradient-echo-planar imaging (EPI)^42^ sequence (75 slices acquired in the axial-oblique plane parallel to the AC–PC line, TR = 1,000 ms, TE = 30 ms, flip angle = 55°, slice thickness = 2 mm, multiband factor = 5, in-plane resolution = 2 mm × 2 mm). Each of the six-task runs and two-resting-state runs was acquired over 6 minutes and 49 seconds, for a total of more than 48 minutes of fMRI data per subject. The total MRI session was less than 90 minutes.

### fMRI Tasks

The first and last functional scans were resting-state runs for which the participants were asked to rest with their eyes open while a fixation cross was displayed. The participants completed each of the six tasks during the remaining runs (described below). Tasks were presented using Psychtoolbox-3^43^ and the task order was counterbalanced across participants. Each task, excluding the Movie Watching task, was prefaced with instructions and practice, which were followed by the opportunity for the participant to ask questions about the task. The participants’ responses were recorded on a two-by-two button box, and a fixation cross was shown between the tasks.

The **Card Guessing**^1-3^ task draws on the reward responsiveness construct. Participants are shown a card with a question mark on it, indicating that they must guess whether the number on the other side of the card is greater or lower than 5. This question mark card is displayed for 1.5 seconds during which time the participant responds (right index finger for <5, right middle finger for >5). As soon as the participant responds, the card “flips over” to reveal the number (between 1 and 4 or between 6 and 9). This number is displayed for 0.5 seconds, after which an arrow (green/up or red/down) is displayed for 0.5 seconds to indicate accuracy (“correct” and “incorrect”, respectively). There is a 1-second inter-trial interval (ITI), and an extra 1-second inter-block interval. The guess accuracy is deterministic. In high-win blocks, participants receive positive feedback (i.e., “correct”) on 70% of trials, and conversely, in high-loss blocks, they receive negative feedback (i.e., “incorrect”) on 70% of trials (Figure 3a). There are 10 trials per block and 10 blocks, with the order of win and loss blocks randomized. The task is preceded by instructions and one block of practice. The task is designed to influence participants’ perceived success and examine their responses under varying reward circumstances.

**Figure 3.**
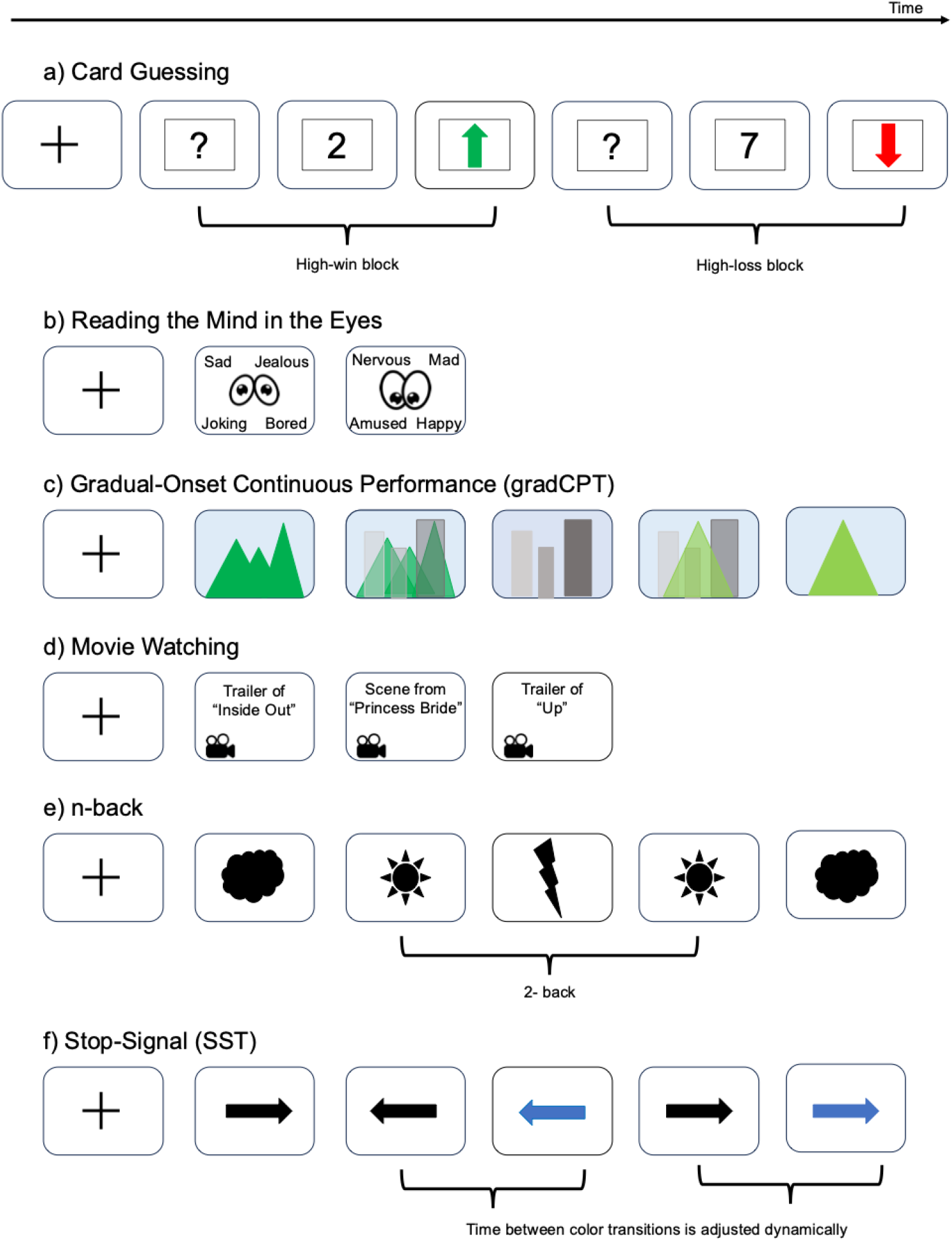
Simplified paradigms of the six in-scanner task runs. All six task runs take the same amount of time (6 minutes and 49 seconds). Each task is described in “fMRI Tasks” under the “Methods” section. A) Card Guessing task. In the high-win block, the green/up arrow (indicating “correct”) is displayed 70% of the time, and in the high-loss block, the red/down arrow (indicating “incorrect”) is displayed 70% of the time. B) Reading the Mind in the Eyes task. The panels here are simplified, while in the actual experiments, photographs of people’s eyes are displayed, not cartoons of eyes. C) Gradual-Onset Continuous Performance task. Here, we showcase the gradual transition between mountains to cities and cities to mountains. D) Movie Watching task. E) n-back task. Here, the panels are simplified to showcase forecast symbols, while in the actual experiments, photographs of human faces and scenery are displayed. F) Stop-Signal task.

The **Reading the Mind in the Eyes**^4^ task draws on the perception and understanding of others constructs. Participants are presented images of a person’s eyes, and must select one out of four adjective choices that best describe the shown person’s mental or emotional state. A paradigm of the task is shown in Figure 3b. Each image is presented for 9.25 seconds or until the subject responds (via button box; each button corresponds to one adjective, with the response mapping counterbalanced across subjects [i.e., up and down are assigned either to index and middle fingers, respectively, or vice versa; right and left are always assigned to right and left hands, respectively], followed by 0.75 seconds of fixation). 36 images are presented. The task is preceded by instructions and one practice trial. The task assesses the ability to interpret subtle social cues and understand others’ thoughts and feelings based on minimal facial information.

The **Gradual-Onset Continuous Performance (gradCPT)**^**5**^ task taps into the attention construct. Participants are presented with a continuous stream of gradually transitioning images, alternating between city and mountain scenes (Figure 3c). They are instructed to press a button in response to images of cities, and to withhold responses when mountains are shown. The task is preceded by instructions and 20 seconds of practice; there is one block, 360 seconds in duration, and 10% target (i.e., mountains). GradCPT is designed to assess sustained attention under conditions of perceptual ambiguity.

The **Movie Watching** task prompts the perception construct. Participants watch three movie clips (Figure 3d) which are each approximately 2 minutes long from three different movies (first the “Inside Out” trailer, second the marriage scene from the “The Princess Bride”, and third the “Up” trailer). Participants are asked to relax and enjoy the movies. No responses are required. The task is designed to influence participants’ visual processing, synchronously stimulating brain states across participants.

The **n-back**^6-8^ task taps into the working memory construct. Participants must decide if a photograph is the same or different than the photograph that came two before; they are instructed to push with their right index finger when the image is different than the one that came two before, but not to push when the image is the same. Images are presented for 1 second, followed by 1 second of fixation. The target (i.e., same image) probability is 10%. There are two blocks (each preceded by a 5-second cue), each with 90 trials. One block uses images of faces, and the other of scenes^44-46^. Block order is randomized for each subject, as is stimulus presentation. The task is preceded by instructions and two short blocks of practice. A simplification of the task is shown in Figure 3e. The task requires participants to maintain information in working memory and compare and update it to the new stimuli.

The **Stop-Signal**^9,10^ task taps into the cognitive control construct. Participants must push a button corresponding to the direction in which a presented arrow is pointing (right index finger for left; right middle finger for right) and must withhold their press when the arrow turns blue (Figure 3f). The arrow is presented for 1.5 seconds, ITI = 0.5 seconds. Signal trial frequency is 0.25, and signal onset is initialized at 250msec; this continuously adjusts based on accuracy (i.e., if a subject correctly withholds a response with a 250msec latency, the latency is increased to 300msec, and so on in a stepwise manner to converge on a probability of stopping of 0.5).

There are 88 trials per block and 2 blocks, with stimulus order randomized within block. Task is preceded by instructions and a short practice block (“block 0”).

### MRI Processing Pipeline

Standard image preprocessing was performed. Skull stripping of the structural scans was done using optiBET^47^, which is an optimized version of the FMRIB’s Software Library (FSL) pipeline^48^. SPM12 was used for motion correction^49^. Nonlinear registration of the MPRAGE to the MNI template was performed using BioImage Suite^50^, and linear registration of the functional to the structural images was done through a combination of FSL and BioImage Suite. The remaining preprocessing steps were performed in BioImage Suite, including regression of mean time courses in white matter, cerebrospinal fluid, and grey matter; high-pass filtering to correct linear, quadratic, and cubic drift; regression of 24 motion parameters; and low-pass filtering (Gaussian filter, *σ* = 1.55)^51^. The processing pipeline code utilized by this study can be accessed at: https://github.com/YaleMRRC/YaleNeuroConnect_ProcessingPipeline.

### Functional connectivity

The Shen268^11^ atlas (and Shen368^12^ atlas) was applied to the preprocessed data, parcellating it into 268 (368) functionally coherent nodes. The mean time courses of each node pair were correlated, and the correlation coefficients were Fisher transformed, generating eight (for the eight in-scanner runs) 268 × 268 (368 × 368) connectivity matrices per subject.

### Neuropsychological and clinical assessment

The MRI scans were followed by a suite of neuropsychological and clinical assessments with both self-reported and examiner-administered components. This test battery was designed to measure a wide range of cognitive, psychiatric, and behavioral constructs across a diverse cohort of participants. To assess clinically relevant symptoms and behavioral phenotypes, we administered seven self-report measures and one psychiatric interview. We report the raw scores, as well as scaled scores that are based on age-normed data. Further information on sub-scales, test type, test length, and reported scores for these assessments can be found in Table 3. The cognitive test battery and the test descriptors can be found in Table 4. All assessments were administered and scored by trained staff, with all of the scoring being done through a two-rater system to ensure consistency and reliability.

## Data Availability

All data produced in this study are available online: https://github.com/YaleMRRC/YaleNeuroConnect.

https://nda.nih.gov/edit_collection.html?id=3276

https://yaleedu-my.sharepoint.com/:f:/g/personal/todd_constable_yale_edu/En99p1wBK6xKj3Zuh2T48_cBVN_oUWEoDlDoVjKhZiMXtQ

https://github.com/YaleMRRC/YaleNeuroConnect_ProcessingPipeline

## Data Records

The YaleNeuroConnect dataset is publicly shared online. The dataset is available in two locations which can be accessed through the YaleNeuroConnect GitHub page (https://github.com/YaleMRRC/YaleNeuroConnect). The structural and functional MRI data, behavioral, and clinical scores are available on the NIMH Data Archive website (https://nda.nih.gov/edit_collection.html?id=3276), and the functional connectomes, mean regions of interest (ROIs), and demographic and diagnostic information are available on the YaleNeuroConnect OneDrive website (https://yaleedu-my.sharepoint.com/:f:/g/personal/todd_constable_yale_edu/En99p1wBK6xKj3Zuh2T48_cBVN_oUWEoDlDoVjKhZiMXtQ).

Access to data on the NIMH website requires an application process, as outlined on the YaleNeuroConnect GitHub page. The YaleNeuroConnect OneDrive site is openly accessible and does not require authorization or password protection.

1. NIMH website (https://nda.nih.gov/edit_collection.html?id=3276):
  – Behavioral measures (n=405)
  – Clinical measures (n=405)
  – Structural MRI (n=405)
  – Raw fMRI runs, 8 runs per participant (n=405)
2. YaleNeuroConnect OneDrive website (https://yaleedu-my.sharepoint.com/:f:/g/personal/todd_constable_yale_edu/En99p1wBK6xKj3Zuh2T48_cBVN_oUWEoDlDoVjKhZiMXtQ).
  – Functional connectomes (n=302)
  – Mean ROIs (n=302)
  – Demographic measures (n=302, n=405)
  – Diagnostic categories (n=302, n=405)
  – Sample data for the fMRI processing pipeline (n=1)
  – README files are included to describe the different folders

### Technical Validation

All registered data were visually examined to ensure whole-brain coverage, adequate registration, and the absence of artifacts or other quality issues. Subjects that did not meet the expected imaging criteria were excluded from the study. N=405 subjects met the expected imaging criteria. Subjects who completed all eight fMRI scans, whose grand mean frame-to-frame displacement was less than 0.15 mm, and whose maximum mean frame-to-frame displacement was less than 0.2 mm were selected for generating the functional connectome data (n=302, additional subjects may be included as the data review progresses). Two participants who were scanned for all or some of the protocols under a slightly shorter scanning time (25 seconds shorter) were not excluded from the dataset, as the missing time segments of these scans were expected.

### Code Availability

The fMRI data pre-processing pipeline code utilized by the YaleNeuroConnect study can be accessed at: https://github.com/YaleMRRC/YaleNeuroConnect_ProcessingPipeline.

The preprocessing pipeline requires the installation of FSL (6.0.1), BioImage Suite (35), and SPM12 to function properly. We provide scripts that convert DICOM files to BIDS format, perform skull stripping, motion correction, and nonlinear registration of individual structural MRIs to a template brain, conduct linear registration of functional scans to the structural MRIs, generate functional connectivity matrices, and apply uniform smoothing. Additionally, data from a single subject at each preprocessing step is provided as a reference.

## Acknowledgements

This work was supported by funding from the NIH (MH121095 to D.S. and R.T.C).

## Author contributions

A.S. wrote the manuscript, with contributions from R.T.C. and S.I., and comments from all authors. A.J.S. prepared the fMRI data pre-processing code. J.A. formatted the data for the NIMH Data Archive website. A.S.G., S.M., J.A., F.T., and C.A.H. collected the dataset, and C.A.H. and S.M. managed the study. G.S., R.K., V.H.S. and S.W.W. supported all clinical aspects of the Yale study. A.S.G. designed the Yale study with support from M.N.S., R.T.C., and D.S.

